# Mass drug administration to reduce malaria incidence in a low-to-moderate endemic setting: short-term impact results from a cluster randomised controlled trial in Senegal

**DOI:** 10.1101/2024.07.17.24310593

**Authors:** El-hadji Ba Konko Ciré, Michelle E. Roh, Abdoulaye Diallo, Tidiane Gadiaga, Amadou Seck, Sylla Thiam, Seynabou Gaye, Ibrahima Diallo, Aminata Colle Lo, Elhadji Diouf, Omar Gallo Ba, Alioune Badara Gueye, Ari Fogelson, Xue Wu, Paul Milligan, Tabitha Kibuka, Moustapha Hama, Erin Eckert, Julie Thwing, Adam Bennett, Roly Gosling, Jimee Hwang, Doudou Sene, Fatou Ba, Bayal Cissé, Katharine Sturm-Ramirez, Michelle S. Hsiang, Jean Louis Ndiaye

**Author notes:** **Corresponding authors:** Michelle E. Roh, UCSF Institute for Global Health Sciences, 550 16th Street, 3rd Floor, San Francisco, CA 94158, USA, El-hadji Konko Ciré Ba, Université Iba Der Thiam de Thiès, Grand Standing, Thiès, BP: A967 Thiès. Contributed equally (El-hadji Ba Konko Ciré, Michelle E. Roh). Contributed equally (Michelle S. Hsiang, Jean Louis Ndiaye).

## Abstract

**Background:** In Africa, the scale-up of malaria control interventions, including seasonal malaria chemoprevention (SMC), has dramatically reduced malaria burden, but progress toward malaria elimination has stalled. We evaluated mass drug administration (MDA) as a strategy to accelerate reductions in malaria incidence in Senegal.

**Methods:** We conducted an open-label, cluster-randomised controlled trial in a low-to-moderate transmission setting of Tambacounda, Senegal. Eligible villages had a population size between 200–800. All villages received pyrethroid-piperonyl butoxide bednets and proactive community case management of malaria at baseline. Sixty villages were randomised 1:1 to either three cycles of MDA with dihydroartemisinin-piperaquine+single-low dose primaquine administered to individuals aged ≥3 months, six-weeks apart starting the third week of June (intervention), or standard-of-care, which included three monthly cycles of SMC with sulfadoxine-pyrimethamine+amodiaquine administered to children aged 3–120 months starting end of July (control). MDA and SMC were delivered door-to-door. The primary outcome was clinical malaria incidence in all ages assessed during the peak transmission season (July-December), the year after intervention. Here, we report safety, coverage, and impact outcomes during the intervention year. The trial is registered at ClinicalTrials.Gov (NCT04864444).

**Findings:** Between June 21, 2021 and October 3, 2021, 6505, 7125, and 7250 participants were administered MDA and 3202, 3174, and 3146 participants were administered SMC across cycles. Coverage of ≥1 dose of MDA drugs was 79%, 82%, and 83% across cycles. During the transmission season of the intervention year, MDA was associated with a 55% [95% CI: 28%– 72%] lower incidence of malaria compared to control (MDA: 93 cases/1000 population; control: 173 cases/1000 population). No serious adverse events were reported in either arm.

**Interpretation:** In low-to-moderate malaria transmission settings with scaled-up malaria control interventions, MDA with dihydroartemisinin-piperaquine+single-low dose primaquine is effective and well-tolerated for reducing malaria incidence. Further analyses will focus on the sustainability of this reduction.

**Funding:** United States President’s Malaria Initiative

**Research in context:** *Evidence before this study:* The current World Health Organization (WHO) guidelines recommend that malaria programmes consider mass drug administration (MDA) for *Plasmodium falciparum* transmission reduction in low-to-very low transmission settings (broadly defined as parasite prevalence <10% or annual malaria incidence of <250 cases per population). In moderate-to-high transmission areas, MDA is recommended for rapid reduction of disease burden, but not for transmission reduction due to the lack of published studies demonstrating its short- or long-term benefits. Among the numerous studies that contributed to this recommendation, five evaluated the antimalarial combination, dihydroartemisinin-piperaquine + single low-dose primaquine. However, none of the studies were conducted in countries implementing seasonal malaria chemoprevention as part of their routine malaria control strategy. On January 23, 2024, we conducted a PubMed search using the following term: “mass drug administration” AND “dihydroartemisinin-piperaquine”. We found one additional cluster randomised controlled trial conducted in a moderate transmission setting of The Gambia (an SMC-implementing country), that evaluated mass drug administration with the antimalarial combination, dihydroartemisinin-piperaquine + ivermectin. This study demonstrated that MDA was associated with a 70% reduction in the odds of PCR-confirmed malaria two months after the last round of MDA. However, given the study demonstrated little evidence on entomological outcomes, authors concluded that much of the observed effect of MDA was likely attributable to the antimalarial efficacy of dihydroartemisinin-piperaquine.

*Added value of this study:* Our study adds to the current evidence base demonstrating the benefits of MDA with dihydroartemisinin-piperaquine + single-dose primaquine on malaria burden reduction and may have impacts on short-term transmission. Combined with The Gambia trial results, our study provides new evidence demonstrating that MDA with dihydroartemisinin-piperaquine can have short-term benefits on transmission in low and moderate transmission settings where malaria transmission is highly seasonal.

*Implications of all the available evidence:* As countries in sub-Sahelian and Sahelian Africa progressively scale-up their malaria control interventions, they will reach a plateau where no further gains can be made. In low and moderate transmission settings, MDA is a well-tolerated and effective intervention for rapidly reducing malaria burden and may have an impact on transmission in the short-term.

## Introduction

Malaria is a major public health concern in Africa. In regions where transmission is highly seasonal, seasonal malaria chemoprevention (SMC) has been widely adopted to prevent morbidity and mortality in children at-risk of severe malaria. SMC involves the monthly administration of sulfadoxine-pyrimethamine + amodiaquine given during the peak transmission season to treat existing parasitaemia and prevent new infections.^1^ Since its initial recommendation by the World Health Organisation (WHO) in 2012, SMC has expanded to 18 African countries, covering 53 million children in 2023.^2^ The scale-up of SMC has been successful, demonstrating reductions in childhood clinical malaria incidence by 60–88% under programmatic conditions.^1,3^

Through high coverage of SMC, strong vector control, and prompt case management, countries in Sahelian and sub-Sahelian Africa have made significant strides in controlling malaria, prompting many to establish new goals for malaria elimination. However, recent progress toward elimination has stalled,^4^ necessitating enhanced coverage of proven core interventions and the consideration of new interventions to rapidly reduce transmission. One promising approach is mass drug administration (MDA), which involves the administration of antimalarials to all individuals in a defined geographic area at a frequency and duration tailored to the local malaria epidemiology and goals. For MDA to have an impact on transmission, high coverage (≥80%)^5,6^ of the target population is needed, which requires an optimized delivery approach and strong community engagement.^7^ Achieving high coverage may be less challenging in countries successfully implementing SMC, as they can leverage their existing infrastructure of door-to-door delivery and community and health system acceptance of chemoprevention.^8,9^

In addition to attaining high coverage, the effectiveness of MDA depends on the type of antimalarial regimen used. In *Plasmodium falciparum*-dominant regions, dihydroartemisinin-piperaquine is an attractive agent for MDA given its good safety profile, long prophylactic period, and relatively low-levels of artemisinin resistance in Africa.^10,11^ However, dihydroartemisinin-piperaquine has no known efficacy against mature gametocytes^12,13^—the parasites responsible for human-to-mosquito transmission. Single low-dose primaquine is a gametocytocidal agent shown to be safe and associated with the near complete prevention of human-to-mosquito transmission.^14,15^ It is likely that the combination of these drugs may confer greater benefits than dihydroartemisinin-piperaquine alone, allowing MDA to have a greater impact on transmission.

Here, we present results from a cluster randomised controlled trial assessing the safety, coverage, and short-term impact of three cycles of MDA with dihydroartemisinin-piperaquine + single low-dose primaquine on *P. falciparum* incidence and prevalence during the intervention year. This study aimed to fill a critical evidence gap regarding the effectiveness of MDA to rapidly reduce malaria burden in a highly seasonal, low-to-moderate transmission setting where malaria control measures have been scaled-up and additional interventions are needed to accelerate malaria elimination.

## Methods

### Study setting

The study was conducted in Tambacounda Health District of southeastern Senegal (**Appendix 1**). The district is comprised of 523 villages with an estimated population size of 297,761 in 2020. In southeastern Senegal, malaria transmission is low-to-moderate (50–200 cases per 1000 population) and highly seasonal, with most cases occurring between July and December. In this region, the national malaria programme (*Programme National de Lutte contre le Paludisme*; PNLP) implements standard malaria control interventions including routine distribution of insecticide-treated nets, malaria case management at health facilities, SMC to children 3–120 months of age (except those with a severe/chronic illness, known hypersensitivity to SMC drugs, and history of antimalarial receipt in the prior three weeks), and proactive community case management of fever through the *Prise en Charge à Domicile Plus* (PECADOM+) model. In the PECADOM+ model, community health workers, known as dispensateur de soins à domiciles (DSDOMs), conduct weekly household visits to identify and treat suspected malaria cases during the malaria transmission season. Despite scale-up of these interventions, progress toward transitioning these zones to pre-elimination status has been slow. Thus, in these areas, the program needs an accelerator intervention to aggressively push the elimination margins and meet the national goal to eliminate malaria by 2030.

### Study design and participants

The study employed a two-arm, open-label, cluster randomised controlled trial design. Sixty villages were randomly selected for participation. Villages were eligible if they had a population size between 200–800; were located within a health post catchment area with an annual malaria incidence of 60–160 cases/1000 population; and had an established PECADOM+ system or the PEACDOM+ model was planned for implementation in the village. Villages were selected so that village centroids were ≥2.5 km apart.

Participant eligibility was assessed prior to each MDA cycle. Residents of intervention villages were eligible for MDA if they were ≥3 months of age and excluded if they reported a severe/chronic illness, had a known hypersensitivity to study drugs, were pregnant (confirmed by urine test), had taken drugs that could influence cardiac function or prolong QTc interval, or received antimalarials in the prior three weeks. Children <2 years of age and breastfeeding women were further excluded from receiving single low-dose primaquine. No SMC was provided during the intervention year in intervention villages.

Written informed consent was obtained before the first MDA cycle and cross-sectional surveys. Parental written informed consent was obtained from participants <18 years of age and written informed assent was obtained from children 13–17 years of age.

Ethical approval of this trial was granted by the Comité National d’Ethique pour la Recherche en Santé (CNERS) of Senegal and the University of California, San Francisco (UCSF) Human Research Protection Program. The U.S. Centers for Disease Control and Prevention approved reliance on UCSF. The trial was registered with ClinicalTrials.gov (NCT04864444) and oversight was provided by an independent data safety monitoring committee and external monitor. An interim safety analysis was conducted after the first MDA cycle.

### Randomisation and masking

Study villages were randomised 1:1 to intervention or control. To ensure adequate balance with respect to specified variables, villages were stratified based on whether PECADOM+ was present at baseline. Then a constrained randomisation approach was undertaken using the following village-level covariates: health post of village, distance to health post, baseline malaria prevalence (assessed through a survey conducted at the end of the pre-intervention transmission season), village population size, and population size of children <10 years. A study investigator (MER) randomly generated intervention assignment. Participants, field team, and investigators were unblinded to allocation assignment. Laboratory technicians were blinded to intervention assignment.

### Procedures

#### Community mobilisation and sensitisation

Upon village selection, the study team held meetings with administrative, health, and religious leaders of Tambacounda Health District to discuss the study aims, planned activities, and receive consent for study implementation. Community sensitisation materials were developed by the study team and implemented by local health staff. In the months prior to MDA, which coincided with the peak COVID-19 pandemic, additional social media campaigns, local community radio announcements, and television advertisements were conducted. Prior to each MDA cycle, town hall meetings and household visits were undertaken to ensure that the community was well-informed.

#### Interventions

In the control arm, the standard-of-care chemoprevention (which consisted of three cycles of SMC with sulfadoxine-pyrimethamine + amodiaquine) was administered to children aged 3-120 months at four-week intervals, initiated at the presumed start of the malaria transmission season. In the intervention arm, three cycles of MDA with dihydroartermisinin-piperaquine + single low-dose primaquine were administered to individuals aged ≥3 months at six-week intervals. To achieve maximal impact on clearing the infectious reservoir,^16,17^ MDA was initiated one month prior to the presumed start of the transmission season (**Figure 1**). Prior to intervention implementation, pyrethroid-piperonyl butoxide (PBO) bednets were distributed door-to-door to all study villages and year-round PECADOM+ was established to monitor malaria incidence.

**Figure 1.**
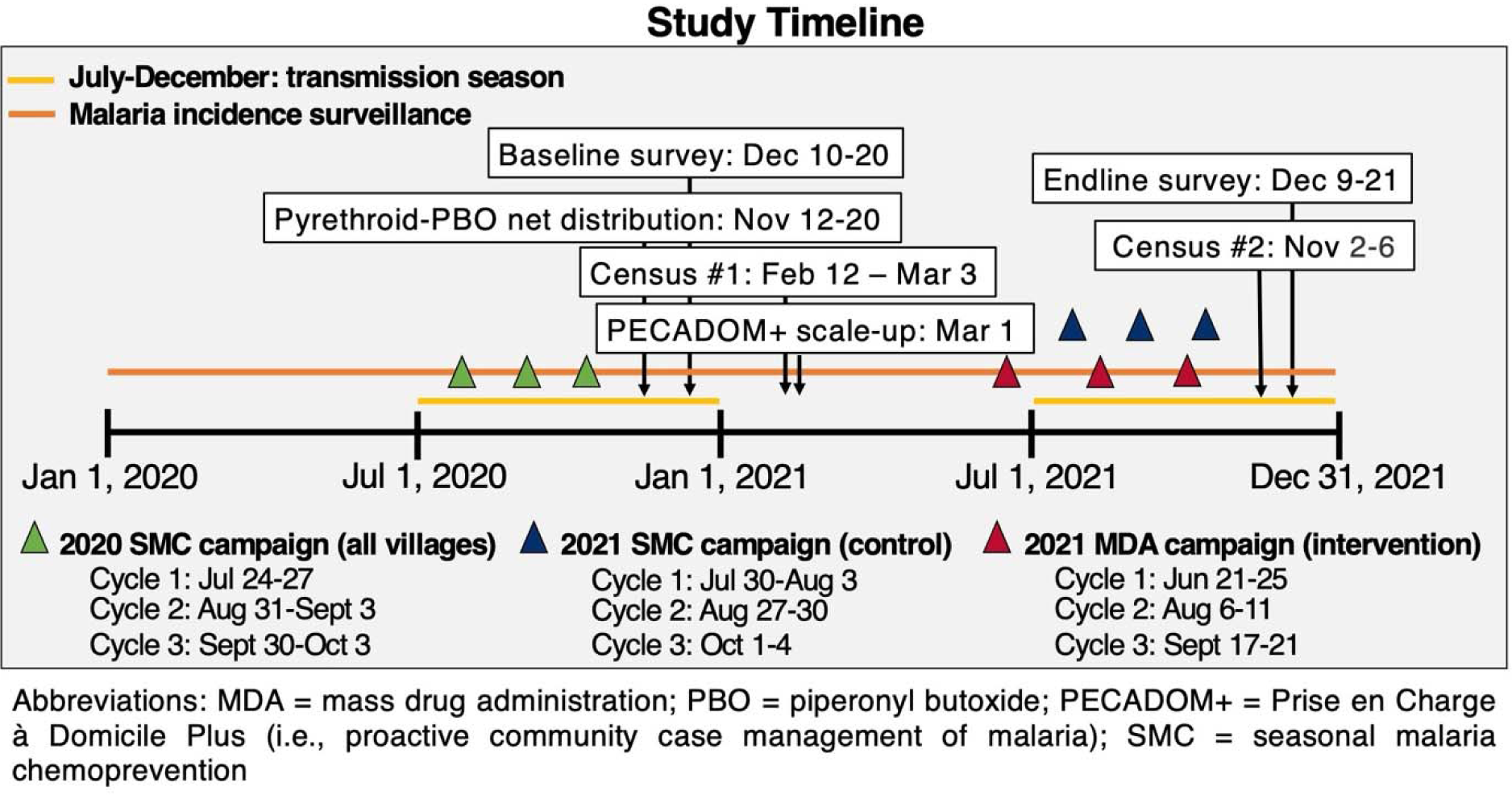
Timeline of study activities

MDA and SMC were delivered door-to-door by DSDOMs via directly observed therapy for all three doses. Study drugs were administered using an age-based dosing strategy per manufacturer’s instructions (**Appendix 2**). For each MDA cycle, dihydroartemisinin-piperaquine (Eurartesim™, Sigma-Tau, Italy) was given for three consecutive days and single low-dose primaquine (Remedica Ltd, Limassol, Cyprus) was given with the first dose of dihydroartemisinin-piperaquine. For each SMC cycle, amodiaquine (Fosun Pharma, Shanghai, China) was given for three consecutive days and sulfadoxine-pyrimethamine (Fosun Pharma, Shanghai, China) was given with the first dose of amodiaquine. For participants who were unable to swallow tablets (e.g., young children), tablets were crushed and mixed in water. If the participant vomited within 30 minutes of administration, the full dose was re-administered. If the re-administered dose was vomited within 30–60 minutes, half the dose was administered.

During drug administration, individuals with symptomatic malaria confirmed by a histidine-rich protein 2-based rapid diagnostic test (RDT) (ParaHIT®-f, ARKRAY Healthcare Pvt Ltd, Surat, India) were treated with artemether-lumefantrine and did not receive study drugs until the subsequent cycle.

#### Malaria surveillance

Malaria cases were captured through health facility and PECADOM+ registries. Suspected cases (i.e., presentation of fever or history of fever in the past 48 hours) were confirmed by RDT. To ensure high-quality capture of incident cases, PECADOM+ was expanded year-round implementation in all study villages and fully scaled-up by March 1, 2021. Data were collected on paper-based registries and abstracted onto electronic databases. Duplicates between registries were removed. Village-level population size was estimated by averaging estimates from two censuses conducted before-and-after intervention implementation (**Figure 1**).

#### Pharmacovigilance

Passive and active pharmacovigilance systems were used to monitor the safety of MDA. For passive surveillance, adverse events (AEs) following drug intake were recorded by study staff or DSDOMs into standardised case report forms. Participants were encouraged to inform local health or study staff if they experienced an AE within one month after drug intake. AEs were graded by a study clinician (e.g., mild, moderate, or severe) and managed free of charge. For active surveillance monitoring, the study staff surveyed 220 random households per study arm on the day after the last dose of study drugs. Households were sampled from every village, proportionate to village population size, such that five households were sampled from villages with <300 residents and ten households from villages with ≥300 residents. In intervention villages, the study team randomly sampled three members per household according to their age group: <5, 5–15, and ≥15 years. In control villages, three household members <10 years of age were randomly sampled. Survey participants were asked if they experienced an AE and to describe the event, including type, onset date, and duration. Severity was graded by a study clinician.

#### Cross-sectional surveys

To determine parasite prevalence, cross-sectional surveys were conducted at the end of the transmission season before-and-after intervention implementation (December 10–20, 2020 and December 9–21, 2021). A two-stage cluster sampling strategy was undertaken to randomly select households and household members from all villages. Participants were asked about their demographic characteristics, malaria prevention measures, and history of fever. Suspected malaria cases were confirmed by RDT and those with a positive test were treated with artemether-lumefantrine. A fingerprick blood sample was taken for microscopy and for dried blood spots (DBS) to confirm parasitaemia by PCR and to genotype drug resistance markers.

#### Laboratory analysis

Microscopy slides and DBS from surveys were transported to Université Iba Der Thiam de Thiès. Slides were stained with 6% Giemsa for 20 min and read by two microscopists. A third reviewer settled discrepant findings. Parasite DNA was extracted from DBS using the Chelex-100 extraction method^18^ and tested for parasitemia by real-time PCR using species-specific primers based on 18s rRNA gene as previously described.^19^ PCR-positive samples were genotyped to assess the presence of point mutations in the *pfK13, pfdhps*, *pfdhfr*, *pfcrt*, and *pfmdr1* genes using high-resolution melting analysis as previously described.^20^

### Study outcomes

The primary outcome of the trial was village-level malaria incidence in the year after intervention implementation. Here, we report the impact of MDA on incidence during the transmission season of the implementation year. Village-level malaria incidence was defined as the number of RDT-confirmed symptomatic malaria cases detected through health post and PECADOM+ surveillance divided by the average village population size obtained from two censuses performed before-and-after intervention implementation (**Figure 1**). Secondary outcomes included parasite prevalence by microscopy and PCR, coverage and safety of MDA, and prevalence of drug resistance markers assessed through cross-sectional surveys.

Coverage was defined according to WHO guidelines^7^. For each chemoprevention campaign, a pre-intervention census was used to generate a registry that determined who would be targeted for each cycle. Data on adherence and dose were recorded for each person and day. The registry was updated throughout the campaign to identify new residents, deaths, and emigrants. Both crude and distributional coverage are reported. Crude coverage was defined as the proportion of residents who received ≥1 dose of study drugs among study residents. Denominator included absences, refusals, and those who did not meet the eligibility criteria. Distribution coverage was defined as the proportion of residents who received ≥1 dose among eligible residents, thereby excluding pregnant women and those with a self-reported illness. Both coverage metrics excluded deaths and emigrants.

### Statistical analysis

All analyses were conducted using Stata version 17·0 or R version 4·2·2. Sample size calculations were based on detecting a 50% relative difference in RDT-confirmed malaria incidence between arms in the year post-intervention. We assumed mass distribution of pyrethroid-PBO bednets, SMC, and scale-up of community case management would cumulatively reduce annual malaria incidence by 50% in the control arm from 100 to 50 cases/1000 population before-and-after intervention implementation. Based on a coefficient of variation of 0.80 and an average cluster size of 250, a sample size of 60 clusters provided 80% power (using a 5% significance level) to detect a 50% relative difference in the MDA arm (intervention effect) using a two-tailed alpha test.

Analyses were carried out using an intention-to-treat approach. Intervention impact on incidence was assessed using mixed-effects Poisson regression with village-level random intercepts. In the unadjusted model, the following indicator variables were included: a treatment indicator that equalled 1 in intervention villages during the implementation year and 0 otherwise and a time variable that equalled 1 during the implementation year and 0 otherwise. Adjusted analyses included covariates used in the stratified constrained randomization scheme. Intervention impact was defined as the percent reduction in incidence between July and December in the intervention arm compared to the control arm (1–incidence rate ratio (IRR_intervention_)*100%). Intervention effects on parasite prevalence were estimated using Poisson regression with robust standard errors. Survey weights accounting for number of households and household size were incorporated into prevalence analyses. Prespecified subgroup analyses were conducted by age group (e.g., ≥ or <10 years of age), DSDOM presence at baseline, and baseline transmission intensity (low versus moderate; low defined as parasite prevalence <10% as defined by WHO^1^) using two-way interaction terms between treatment and subgroup variables.

## Results

Between September 1, 2020 and October 25, 2020, 523 villages in the study area were geolocated and screened for eligibility, and 111 met the inclusion criteria (**Figure 2**). Of these, 60 villages were randomly selected allowing for a ≥2.5 km distance between village centroids and randomised to intervention or control. Village-level factors included in the constrained randomisation were balanced across arms (**Table 1**). Overall, coverage of pyrethroid-PBO bednets was high (98%) and similar between arms. In the pre-intervention year, 81% of children <10 years of age reported receiving the most recent cycle of SMC and 71% reported receiving all three cycles. Twenty percent reported sleeping away from their home in the past 15 days.

**Figure 2.**
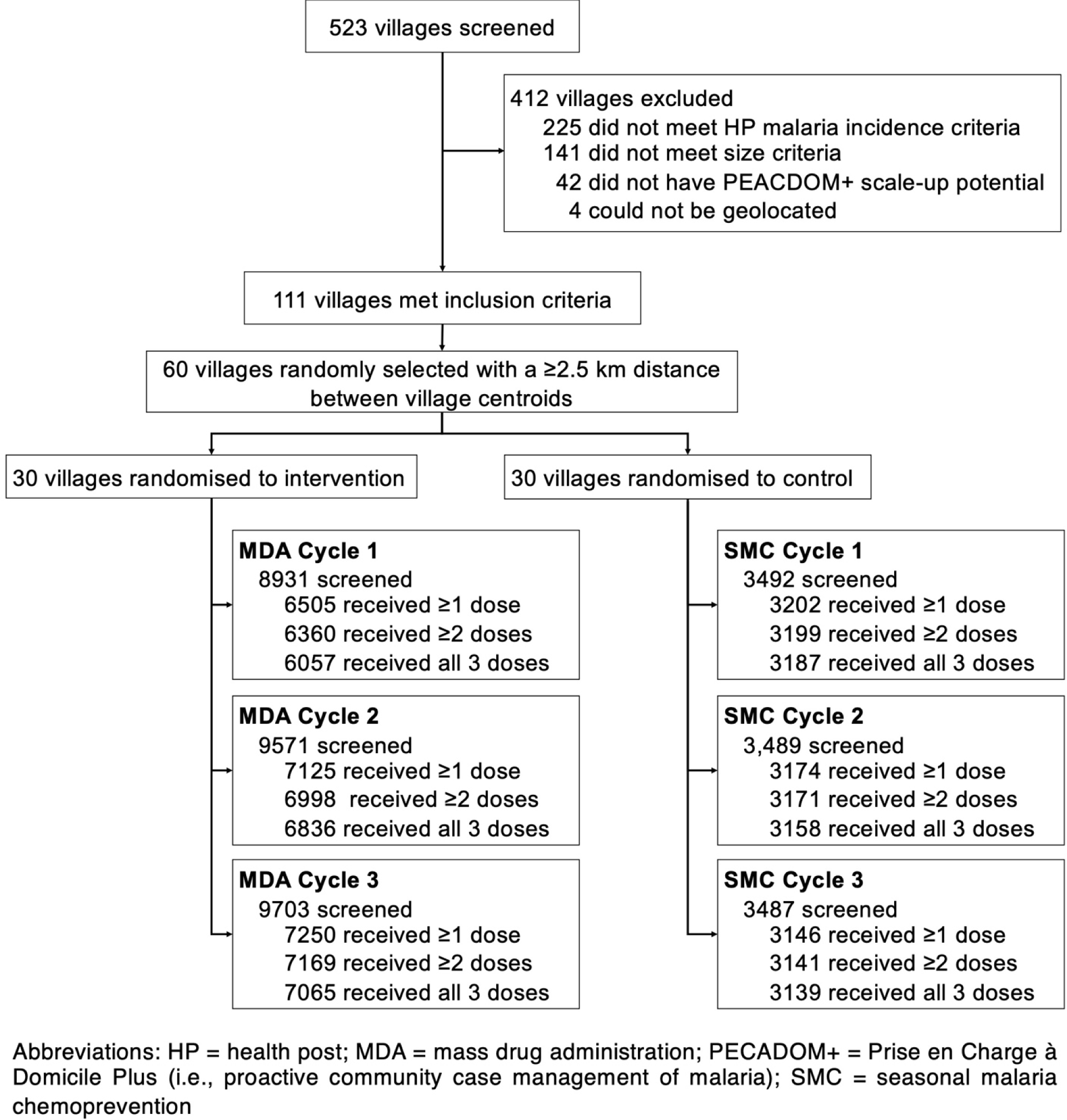
Trial profile

**Table 1.**
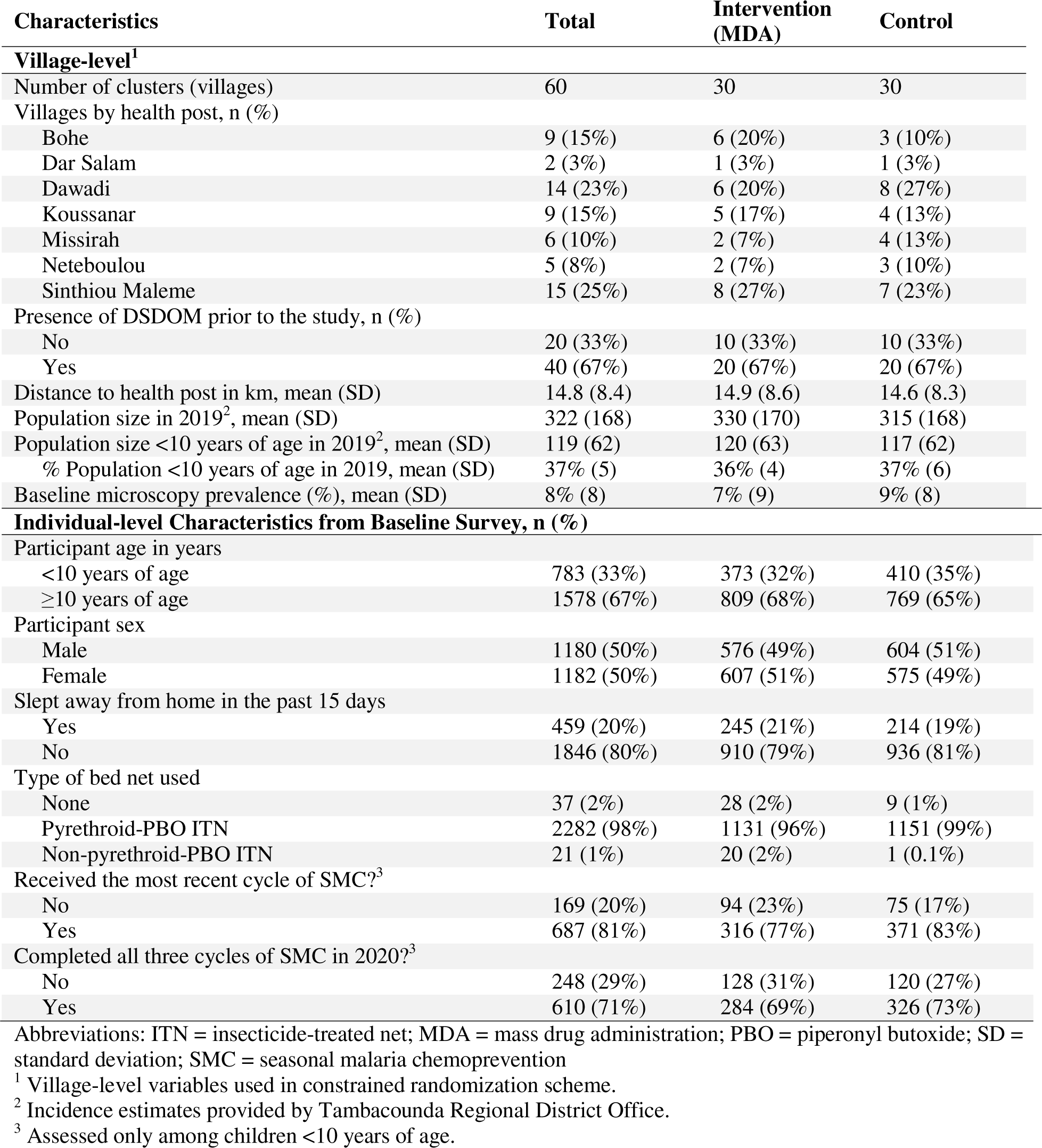
Trial profile.

In the intervention arm, 8931, 9571, and 9703 residents were screened for the first, second, and third cycle of the MDA campaign. Distribution coverage of single low-dose primaquine (where the denominator excluded ineligible residents) was 79% (6286/7992), 82% (6949/8462), and 84% (7199/8575) across the three cycles. Distribution coverage of ≥1 dose of dihydroartemisinin-piperaquine among the eligible individuals was 79% (6505/8229), 82% (7125/8673), and 83% (7250/8690) in the first, second, and third cycles. Distribution coverage of all three doses of dihydroartemisinin-piperaquine was 74%, 79%, and 81% across cycles.

Distribution coverage was higher in those <10 years compared to ≥10 years (distribution coverage of ≥1 dose was 85%, 86%, and 87% in <10 years across cycles, and 75%, 80%, and 81% in ≥10 years across cycles). Crude coverage of ≥1 dose of dihydroartemisinin-piperaquine was 73%, 74%, and 75% across cycles. By village, distribution coverage ranged from 58%–97%; 50%, 67%, and 70% of intervention villages reached the WHO target coverage of ≥80% in the first, second, and third cycles, respectively (**Appendix 3**). The major reasons for non-participation were absence (range: 14%–21%) and illness (range: 5%–7%) (**Appendix 4**).

Absences were similar between males and females (1⋅12:1 ratio) and in age to those who received MDA (16 years [interquartile range (IQR): 7–26] versus 13 years [IQR: 6–27]). Refusals were rare (1%–2%) and mostly among males (70%) with a median age of 22 years [IQR: 15–30].

In the control arm, 3492, 3489, and 3487 children aged 3–120 months were screened for the first, second, and third SMC cycle. Distribution coverage of ≥1 dose of SMC drugs was 93% (3202/3457), 92% (3174/3442), and 92% (3146/3434) across cycles. Distribution coverage for all three SMC doses was 92%, 92%, and 91% across cycles. Crude coverage of ≥1 dose was 92%, 91%, and 90% across cycles. By village, distribution coverage ranged from 62%–100%; 93% of SMC villages reached ≥80% coverage across all cycles (**Appendix 3**). Major reasons for non-receipt of SMC were absence (7%) and illness (1%–2%) (**Appendix 4**). Refusals of SMC were low across cycles (≤1%).

During the pre-intervention transmission season (July-December 2020), malaria incidence was 181 and 204 cases/1000 population in the intervention and control arms, respectively (**Table 2**). In the transmission season of the intervention year (July-December 2021), malaria incidence reduced to 93 cases/1000 population in the intervention arm and to 173 cases/1000 population in the control arm (**Table 2**; **Figure 3**). The unadjusted intervention effect of MDA was 52% [95% CI: 21%, 71%]. The adjusted intervention effect, which accounted for variables included in the constrained randomisation was 55% [95% CI: 28%, 72%].

**Figure 3.**
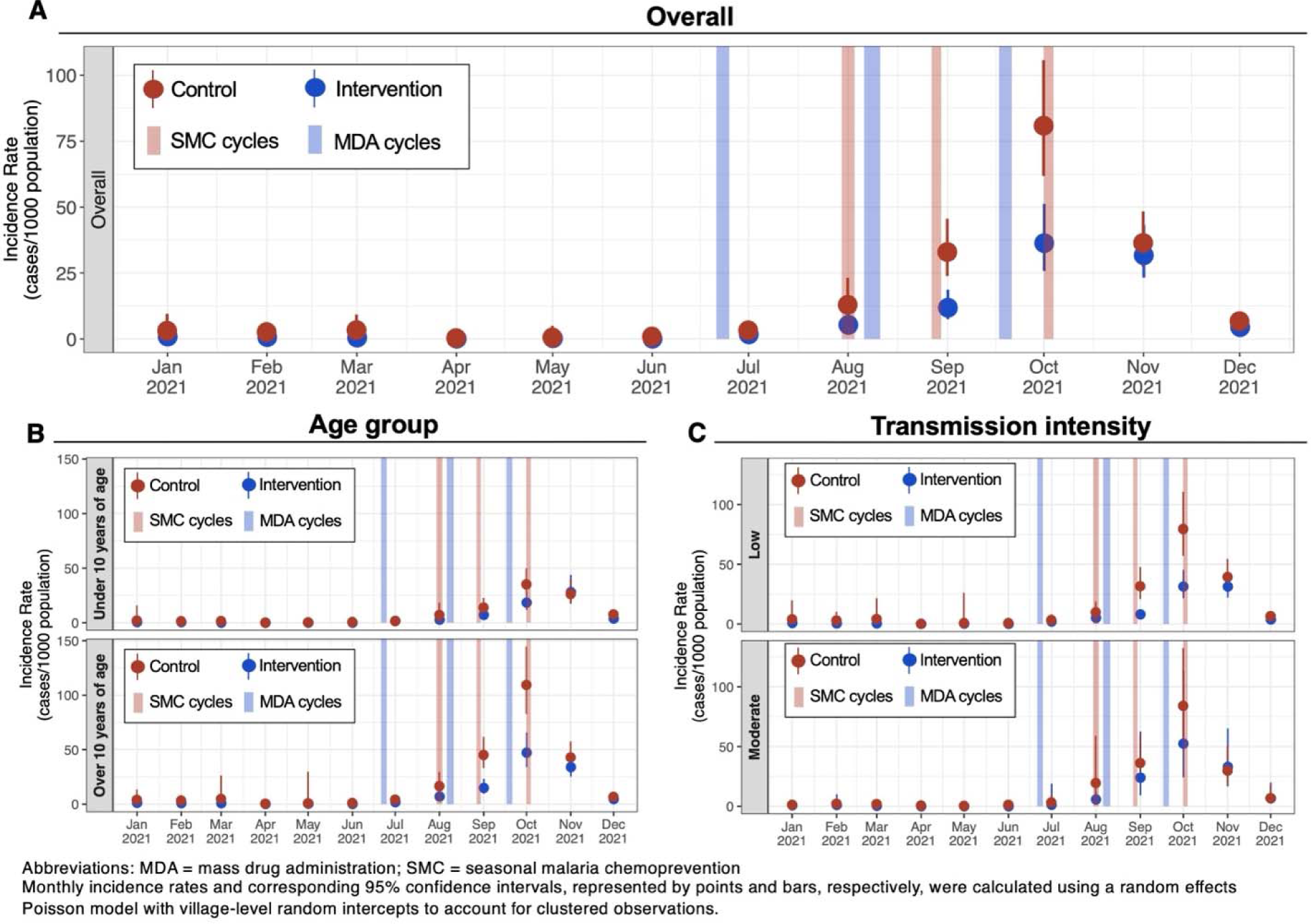
Graphical representation of village-level monthly incidence of symptomatic malaria during the intervention implementation year by intervention group (A) and stratified by age group (B), and baseline transmission intensity (C).

**Table 2.** RDT-confirmed malaria incidence and intervention effectiveness estimates during peak transmission season (July-December)

| Study outcomes | Average incidence per 1,000 (events / population) |  |  |  | Intervention effect,<br>1-ratio of IRRs [95% CI]<sup>1</sup> |  |
| --- | --- | --- | --- | --- | --- | --- |
|  | Intervention (MDA) |  | Control |  | Unadjusted | Adjusted<sup>2</sup> |
|  | 2020 | 2021 | 2020 | 2021 |  |  |
| Primary outcome |  |  |  |  |  |  |
| Incidence among all ages | 181<br>(2067/10745) | 93<br>(924/10745) | 204<br>(1883/9641) | 173<br>(1751/9641) | 52% [21%, 71%] | 55% [28%, 72%] |
| <10 years old | 133<br>(514/4091) | 84<br>(287/4091) | 154<br>(515/3598) | 120<br>(463/3598) | 43% [3%, 66%] | 46% [10%, 67%] |
| ≥10 years and older | 213<br>(1553/6637) | 100<br>(633/6637) | 236<br>(1368/5991) | 206<br>(1287/5991) | 55% [27%, 73%] | 58% [33%, 74%] |
| Subgroup analyses |  |  |  |  |  |  |
| Among children <10 years of age |  |  |  |  |  |  |
| <5 years of age | 102<br>(163/1939) | 69<br>(115/1939) | 133<br>(208/1723) | 93<br>(166/1723) | 29% [-35%, 62%] | 31% [-25%, 62%] |
| 5 to <10 years of age | 85<br>(351/2152) | 61<br>(172/2152) | 118<br>(307/1875) | 85<br>(297/1875) | 43% [-4%, 69%] | 45% [4%, 68%] |
| Baseline transmission intensity<sup>3</sup> |  |  |  |  |  |  |
| Low (<10%) | 172<br>(1508/8316) | 83<br>(658/8316) | 228<br>(1385/6463) | 171<br>(1085/6463) | 53% [25%, 71%] | 55% [31%, 71%] |
| Moderate (≥10%) | 214<br>(559/2429) | 125<br>(266/2429) | 148<br>(498/3178) | 178<br>(666/3178) | 49% [-48%, 82%] | 52% [-19%, 81%] |

Abbreviations: IRR = incidence rate ratio; MDA = mass drug administration
<sup>1</sup> Intervention effect estimated using mixed effects Poisson regression with village-level random intercepts.
<sup>2</sup> Adjusted for DSDOM at baseline, village population size of children <10 years of age, distance from health post, baseline parasite prevalence by microscopy, and village population size in 2019, and presence of DSDOM at time of case detection.
<sup>3</sup> Transmission intensity was defined as baseline village-level microscopy-confirmed parasite prevalence of <10% for “Low” transmission clusters and ≥10% for “Moderate” transmission clusters. 23 clusters from the intervention arm and 21 clusters from the control arm contributed to “Low” category; 7 clusters in intervention arm and 9 clusters in control arm contributed to “Moderate” category.

Pre-specified subgroup analyses showed evidence of an interaction by age group; the adjusted intervention effect was 58% [95% CI: 33%, 74%] in the ≥10 years age group and 46% [95% CI: 10%, 67%] in the <10 years age group (p_interaction_=0·016) (**Table 2**; **Figure 3**). The impact of MDA on malaria incidence was similar in low and moderate transmission settings defined as microscopy parasite prevalence of <10% versus ≥10% (adjusted intervention effect=55% [31%, 71%] versus 52% [-19%, 81%]; p_interaction_=0·88) (**Table 2**; **Figure 3**).

By microscopy, parasite prevalence decreased between 2020 and 2021, from 6·1% to 1·8% in the intervention arm and from 6·7% to 4·7% in the control arm (adjusted intervention effect=62% [95% CI: 22%, 80%]) (**Table 3**). The impact of MDA differed between the <10 and ≥10 years age group (adjusted intervention effect=76% [95% CI: 42%, 90%] versus 51% [95%% CI: -14%, 79%]), but this finding did not reach statistical significance (p_interaction_=0·17). By PCR, parasite prevalence decreased between 2020 and 2021, from 17·9% to 4·5% in the intervention arm and 19·9% to 8·3% in the control arm (adjusted intervention effect=47% [95% CI: 3%, 71%]). The effect of MDA differed between the <10 and ≥10 years age groups: adjusted intervention effect=71% [95% CI: 35%, 87%] and 33% [95% CI: -27%, 65%], respectively (p_interaction_=0·050). The impact of MDA on microscopy- or PCR-confirmed parasite prevalence did not significantly differ between low and moderate transmission settings (p_interaction_ for microscopy-confirmed prevalence=0·73, p_interaction_ for PCR-confirmed prevalence=0·73) (**Table 3**).

**Table 3.** Secondary outcomes of parasite prevalence.

| Outcomes | Mean prevalence, % [95% CI]<sup>1</sup> |  |  |  | Intervention effect, 1-ratio of PRs [95% CI]<sup>1</sup> |  |
| --- | --- | --- | --- | --- | --- | --- |
|  | Intervention (MDA) |  | Control |  | Unadjusted | Adjusted<sup>2</sup> |
|  | 2020 | 2021 | 2020 | 2021 |  |  |
| Microscopy-detected infections |  |  |  |  |  |  |
| All ages | 6.1%<br>[2.8%, 9.4%] | 1.8%<br>[0.8%, 2.9%] | 6.7%<br>[4.0%, 9.4%] | 4.7%<br>[2.5%, 6.9%] | 61% [21%, 81%] | 62% [22%, 80%] |
| < 10 years old | 8.2%<br>[2.7%, 13.7%] | 1.3%<br>[0.2%, 2.4%] | 7.3%<br>[3.1%, 11.6%] | 4.5%<br>[2.0%, 7.0%] | 76% [41%, 90%] | 76% [42%, 90%] |
| ≥ 10 years and older | 5.1%<br>[2.6%, 7.7%] | 2.2%<br>[0.7%, 3.6%] | 6.4%<br>[3.9%, 8.8%] | 4.8%<br>[1.8%, 7.9%] | 50% [-16%, 79%] | 51% [-14%, 79%] |
| Baseline transmission intensity<sup>3</sup> |  |  |  |  |  |  |
| Low (<10%) | 2.6%<br>[1.3%, 3.9%] | 1.0%<br>[0.2%, 1.8%] | 3.4%<br>[2.1%, 4.8%] | 4.3%<br>[1.4%, 7.2%] | 63% [-1%, 86%] | 62% [0%, 86%] |
| Moderate (≥10%) | 18.8%<br>[9.4%, 28.2%] | 5.0%<br>[2.1%, 7.8%] | 17.1%<br>[12.5%, 21.8%] | 6.0%<br>[3.4%, 8.6%] | 54% [18%, 73%] | 56% [22%, 75%] |
| PCR-detected infections |  |  |  |  |  |  |
| All ages | 17.9%<br>[11.3%, 24.4%] | 4.5%<br>[2.5%, 6.4%] | 19.9%<br>[11.8%, 28.1%] | 8.3%<br>[4.8%, 11.7%] | 46% [-3%, 70%] | 47% [3%, 71%] |
| < 10 years old | 18.6%<br>[10.9%, 26.4%] | 2.3%<br>[0.6%, 3.9%] | 19.6%<br>[9.9%, 29.4%] | 5.1%<br>[2.2%, 8.0%] | 71% [34%, 87%] | 71% [35%, 87%] |
| ≥ 10 years and older | 17.5%<br>[11.0%, 24.0%] | 5.8%<br>[2.9%, 8.6%] | 20.1%<br>[11.7%, 28.5%] | 10.2%<br>[5.2%, 15.3%] | 33% [-27%, 64%] | 33% [-27%, 65%] |
| Baseline transmission intensity<sup>3</sup> |  |  |  |  |  |  |
| Low (<10%) | 14.0%<br>[7.9%, 20.1%] | 3.0%<br>[1.0%, 5.0%] | 14.3%<br>[6.3%, 22.3%] | 4.7%<br>[2.4%, 6.9%] | 48% [-4%, 74%] | 47% [-7%, 74%] |
| Moderate (≥10%) | 32.0%<br>[11.4%, 52.5%] | 9.9%<br>[6.1%, 13.7%] | 38.0%<br>[21.7%, 54.4%] | 18.7%<br>[12.2%, 25.3%] | 37% [2%, 59%] | 40% [9%, 60%] |

Abbreviations: MDA = mass drug administration; PCR = polymerase chain reaction; PR = prevalence ratio
<sup>1</sup> Intervention effect estimated using Poisson regression with robust standard errors. Survey weights incorporated to account for two-stage sampling design (by household and household size).
<sup>2</sup> Adjusted for variables included in constrained randomisation: presence of DSDOM at baseline, village population size of children <10 years of age, distance from health post, and village population size in 2019.
<sup>3</sup> Transmission intensity was defined as baseline village-level microscopy-confirmed parasite prevalence of <10% for “Low” transmission clusters and ≥10% for “Moderate” transmission clusters. 23 clusters from the intervention arm and 21 clusters from the control arm contributed to “Low” category; 7 clusters in intervention arm and 9 clusters in control arm contributed to “Moderate” category.

In both passive and active pharmacovigilance systems, the frequency of AEs reduced with each subsequent cycle and no serious adverse events (SAEs) were detected in either arm (**Appendix 5**; **Table 4**). Through passive surveillance, 129 AEs were observed in 67/20 887 (0·003%) participants of the intervention arm and four AEs were observed in 2/9524 (0·0002%) participants of the control arm; all were mild (**Appendix 5**). Both the frequency and proportion of participants who experienced an AE were higher in the intervention arm compared to the control arm (p<0·0001). The most common AEs found in the intervention arm were gastrointestinal issues (45·7%), headaches (25·6%), and fever (17·1%).

**Table 4.** Safety outcomes monitored through active surveillance.

| Active Surveillance | Intervention (MDA) | Control (SMC) | p-value<sup>1</sup> |
| --- | --- | --- | --- |
| **Among all participants surveyed** |  |  |  |
| Participants with any AE, n/N (%) |  |  |  |
| All cycles | 260/1903 (13.7%) | 152/1616 (9.4%) | <0.0001 |
| First cycle | 124/619 (20.0%) | 93/569 (16.3%) | 0.10 |
| Second cycle | 94/628 (15.0%) | 51/563 (9.1%) | 0.002 |
| Third cycle | 42/656 (6.4%) | 8/484 (1.7%) | <0.0001 |
| Frequency of adverse events, n |  |  |  |
| All cycles | 498 | 310 | -- |
| First cycle | 229 | 175 | -- |
| Second cycle | 181 | 119 | -- |
| Third cycle | 88 | 16 | -- |
| Most common adverse events, n/N (%) |  |  |  |
| Gastrointestinal issues<sup>2</sup> | 205/498 (41.2%) | 144/310 (46.5%) | 0.16 |
| Fever | 113/498 (22.7%) | 109/310 (35.2%) | <0.001 |
| Headaches | 59/498 (11.8%) | 20/310 (6.5%) | 0.017 |
| Drowsiness | 26/498 (5.2%) | 8/310 (2.6%) | 0.102 |
| Other | 95/498 (19.1%) | 29/310 (9.4%) | <0.001 |
| Severity of AEs, n/N (%) |  |  |  |
| Mild | 319/498 (64.1%) | 178/310 (57.4%) | 0.059 |
| Moderate | 164/498 (32.9%) | 118/310 (38.1%) | 0.14 |
| Severe | 15/498 (3.0%) | 14/310 (4.5%) | 0.26 |
| Any serious AE, n/N (%) | 0/498 (0%) | 0/310 (0%) | -- |
| **Among children <10 years of age** |  |  |  |
| Participants with any AE, n/N (%) |  |  |  |
| All cycles | 85/841 (10.1%) | 145/1544 (9.4%) | 0.57 |
| First cycle | 36/259 (13.9%) | 90/547 (16.5%) | 0.35 |
| Second cycle | 33/271 (12.2%) | 47/529 (8.9%) | 0.14 |
| Third cycle | 16/311 (5.1%) | 8/468 (1.7%) | 0.007 |
| Frequency of adverse events, n |  |  |  |
| All cycles | 176 | 296 | -- |
| First cycle | 68 | 170 | -- |
| Second cycle | 67 | 110 | -- |
| Third cycle | 41 | 16 | -- |
| Most common adverse events, n/N (%) |  |  |  |
| Gastrointestinal issues<sup>2</sup> | 87/176 (49.4%) | 136/296 (45.9%) | 0.523 |
| Fever | 50/176 (28.4%) | 106/296 (35.8%) | 0.121 |
| Headache | 10/176 (5.7%) | 19/296 (6.4%) | 0.901 |
| Drowsiness | 2/176 (1.1%) | 8/296 (2.7%) | 0.334 |
| Other | 27/176 (15.3%) | 27/296 (9.1%) | 0.0057 |
| Severity of AEs, n/N (%) |  |  |  |
| Mild | 102/176 (58.0%) | 169/296 (57.1%) | 0.86 |
| Moderate | 69/176 (39.2%) | 115/296 (38.9%) | 0.94 |
| Severe | 5/176 (2.8%) | 12/296 (4.1%) | 0.49 |
| Any serious AE, n/N (%) | 0/176 (0%) | 0/296 (0%) | -- |

Abbreviations: AE = adverse event
<sup>1</sup> P-value computed using Chi-squared test or Fisher's exact test (if frequency of any cell value was <5)
<sup>2</sup> Defined as one of the following AEs: nausea, vomiting, diarrhoea, abdominal pain, or loss of appetite.

Through active surveillance, more participants of the intervention arm reported an AE compared to the control arm (13·7% versus 9·4%; p<0·0001) (**Table 4**). Among children aged <10 years, the proportion of reported AEs were similar between arms (10·1% versus 9·4%; p=0·57). In the intervention arm, the most common AEs were gastrointestinal issues (41·2%), fever (22·7%), and headaches (11·8%). Of the 498 AEs reported in the intervention arm, 64·1% were mild, 32·9% were moderate, and 3·0% were severe. Severe AEs included fever (n=7), headache (n=4), drowsiness (n=1), vomiting (n=1), diarrhoea (n=1), and loss of appetite (n=1). All AEs appeared three hours after drug intake, resolved within 72 hours, and did not require hospitalization. No cases of anaemia were found through either passive or active surveillance.

Of the 597 PCR-positive samples collected from surveys, 433 were successfully genotyped to determine molecular markers of antimalarial resistance (**Appendix 6**). Mutations associated with intermediate resistance to SP (PfDHFR N51I, C59R, S108N and PfDHPS A437G) were seen at high proportions (range: 58%–100%). PfDHFR I164L was seen at proportions of 2%–12%. None of the studied mutations differed in proportions between arms or time periods. There was no evidence of PfDHPS K540E, PfDHPS A581G, or PfK13 C580Y mutations.

## Discussion

We conducted a cluster randomised controlled trial evaluating the safety, coverage, and short-term effectiveness of three rounds of MDA with dihydroartemisinin-piperaquine + single low-dose primaquine in a low-to-moderate transmission setting of Senegal where malaria control interventions were scaled-up through pyrethroid-PBO bednet distribution, three monthly SMC cycles, and expansion of year-round proactive community case management of malaria. During the trial intervention year, malaria incidence and parasite prevalence reduced in both arms, likely due in part to pyrethroid-PBO bednet distribution. In intervention villages, MDA was associated with a 55% and 62% reduction in malaria incidence and microscopy-confirmed parasite prevalence. Subgroup analyses showed that MDA had a substantial impact on incidence and parasite prevalence in those <10 and ≥10 years of age and in both low and moderate transmission settings. Overall, MDA was well-tolerated; most AEs were mild or moderate and no cases of anaemia or SAEs were observed.

While the trial was not designed to separately estimate MDA’s effect on transmission versus its direct, prophylactic effect, there was some evidence to suggest that MDA indirectly affected transmission. First, subgroup analyses demonstrated that among children under ten, MDA was associated with a 46% reduction in incidence as compared to children under ten in the control arm who received SMC. We expect that this reduction is unlikely by differences in the prophylactic effects of these drugs given we expect dihydroartemisinin-piperaquine and sulfadoxine-pyrimethamine + amodiaquine to have similar protective efficacies,^11^ MDA covered less of the transmission season, and MDA had slightly lower coverage than SMC in under tens. Moreover, MDA’s impact on parasite prevalence further supports this reasoning, given the endline survey was conducted more than two months after the last cycle of drug administration when the prophylactic effects of both MDA and SMC drugs would have waned.

Our study has several important findings/caveats. First, our monthly incidence analyses (**Figure 4**) revealed that SMC should have started one month later and both chemoprevention campaigns should have been extended to four cycles to cover the entire transmission season. If this had this been done, the impact of MDA in children under tens might not have been as substantial. Second, our findings may not be generalisable to most settings deploying SMC where the malaria burden is mostly moderate-to-high. While our study found MDA was associated with reductions in malaria burden and short-term transmission, evidence of its sustained benefit and cost-effectiveness is needed before considering MDA as an intermediate intervention to accelerate malaria elimination.

The WHO currently recommends MDA for transmission reduction in very low-to-low transmission settings (i.e., parasite prevalence of <10% or incidence <250 cases/1000 population). This recommendation is based on evidence from eight cluster randomised controlled trials which found MDA can have a substantial, but short-term impact in these settings.^1,21,22^ However, MDA is not recommended for transmission reduction in moderate-to-high transmission settings, based on evidence from two cluster randomised trials and two non-randomized studies which did not show significant short- or long-term impacts on prevalence or incidence. Our study provides new evidence indicating MDA may have a short-term impact on transmission in both low and moderate transmission strata. Our findings are consistent with a recent trial conducted in a moderate transmission setting of The Gambia^23^ which demonstrated a 70% reduction in the odds of PCR-confirmed infection two months after the last MDA cycle. Evidence from these two studies should be considered when determining future recommendations in moderate transmission settings.

There were several strengths of the study, including our large sample size, rigorous monitoring of safety, and achievement of high coverage. By the final MDA cycle, 70% (21/30) of villages reached the WHO target for MDA coverage (≥80%)^7^, despite significant operational challenges in door-to-door drug administration amidst the peak of the global COVID-19 pandemic. The high coverage observed in our study was likely attributable to community acceptance of annual SMC campaigns, established infrastructure of community case management, support from key administrative and health authorities, and repeated community sensitisation campaigns. However, absences were common during MDA cycles (14-21%), especially among adolescents and young adults who are generally asymptomatic and known to be important drivers of transmission.^24^ Thus, in order to sustain gains made, future MDA campaigns will need to consider additional strategies to reach these groups in order to prevent malaria importation and potential resurgence after MDA.

Our study had a few limitations. First, as explained earlier, SMC was not optimally timed, making it difficult to draw direct comparisons between SMC and MDA for burden reduction in children. Second, baseline incidence was likely non-differentially mismeasured in villages where DSDOMs were absent at the start of the study (33% of villages). While the stratified randomization scheme balanced this factor between arms, this may have biased our effect estimates toward the null (see **Appendix 7** for analyses restricted to villages with prior PECADOM+). Third, given baseline incidence was likely mismeasured, subgroup analyses by baseline malaria incidence were not reported. Fourth, parasitological confirmation of malaria incidence relied on RDT which could have resulted in false positives potentially biasing our effect estimates toward the null. Fifth, MDA was conducted for only one year and additional cycles may have been needed to reach pre-elimination status (i.e., incidence <5 cases/1000 population). Finally, the current analyses only evaluated short-term impact. After discontinuation of MDA, there is potential for malaria to rebound, and analyses of longer-term impact and cost-effectiveness from an additional year of follow-up of these cohorts are forthcoming which will help to determine the sustainability of MDA.

In low-to-moderate malaria transmission settings, where transmission is seasonal and coverage of standard malaria control interventions is high, we demonstrated that three cycles of MDA with dihydroartemisinin-piperaquine + single-low dose primaquine could safely and rapidly reduce malaria burden and potentially have a short-term impact on transmission. However, future work investigating whether the impact of MDA can be sustained, and its cost-effectiveness is needed to inform countries of the potential of MDA as an intervention to accelerate toward elimination.

## Data Availability

De-identified data may be available upon reasonable request and approval from Principal Investigators via a data sharing agreement.

## Acknowledgements

We are grateful to the community of Tambacounda Health District for their participation in the study. We thank the health workers, district health staff, the PNLP, and research team at Université Iba Der Thiam de Thiès for their contributions in implementing the study and to PMI Vectorlink for conducting pyrethroid-PBO bednet distribution and the early village population census. We also thank our data safety and monitoring board members Prof Umberto D’Alessandro (chair), Prof Jean Guadart, Prof Menno R Smit, Dr Houda Sefiani, and Dr Thomas J Peto for their guidance and oversight of the study. We would also like to thank Drs Meera Venkatesan, Meghna Desai, and Leah Moriarty for their contributions in the early protocol development and study planning, Dr Mame Birame Diouf for his initial coordination and strategic engagement of in-country partners, Drs Jean Biyik and Abdoulaye Diop for their support in coordinating the pyrethroid-PBO bednet distribution, and Dr Hannah Slater for providing the mathematical modelling analysis used for our sample size calculations.

## Contributions

JH conceived of the study and wrote the first draft of the protocol. RG, JLN, MER, AB, KSR, JT, EBKC, FB, ST, ID, and ABG contributed to the protocol and approved of the final version. EBKC, AD, TG, AS, ST, SG, ACL, ED, ID, OGB, and JLN implemented the study. JLN, EBKC, AD, TG, ABG, ST, KSR, MER, MSH, TK, MH, EE, RG, DS, and FB provided oversight of the study. ACL conducted laboratory analysis of samples. The statistical analysis plan was developed by MSH, PM, AF, MER, AB, EBKC, and JLN and data were analysed by AF with XW, MER, AS, and EBKC. MER, EBKC, AF, XW, PM, AB, JH, KSR, MSH, and JLN interpreted the results. MER, EBKC, MSH, and JLN wrote the first draft of the manuscript. PM, JH, AF, and KSR provided additional inputs to the writing. All authors reviewed and approved the final manuscript before submission.

