## Supplementary Appendix for "Mass drug administration to reduce malaria incidence in a low-to-moderate endemic setting: short-term impact results from a cluster randomised controlled trial in Senegal"

**Appendix 1.** Map of study site

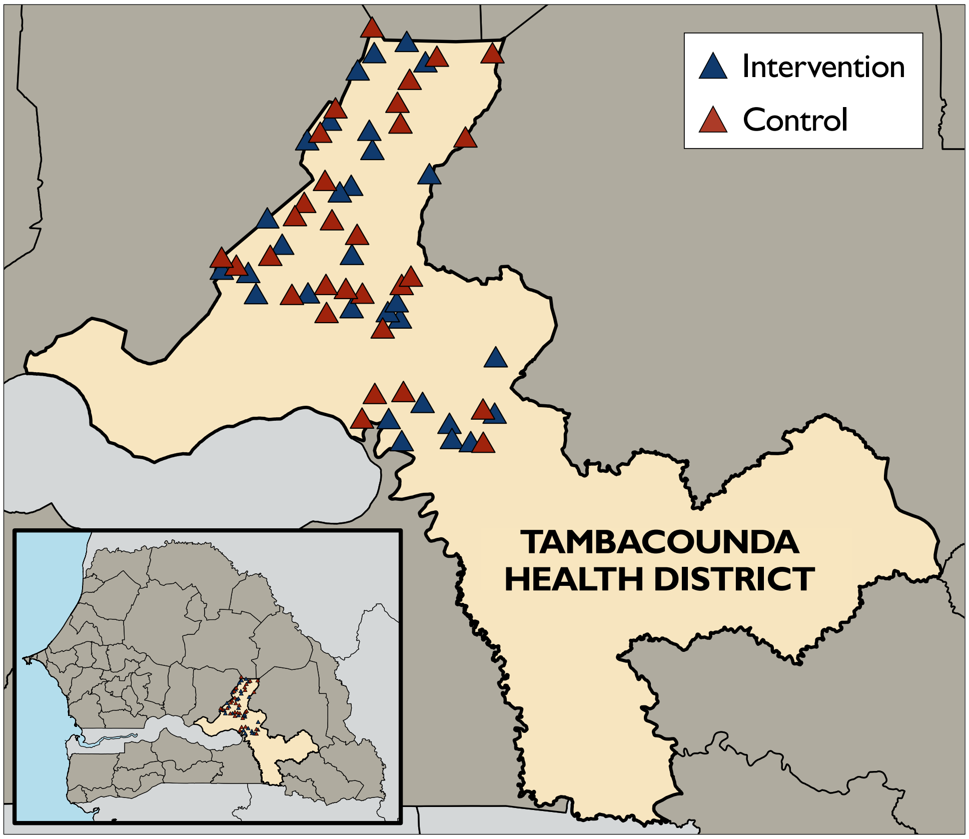

**Appendix 2.** Dosing regimens for dihydroartemisinin-piperaquine, single-low dose primaquine, and sulfadoxine-pyrimethamine + amodiaquine

**Mass drug administration (intervention arm):**

| **Dihydroartemisinin-piperaquine** | | | |
|---|---|---|---|
| Age | Number of tablets containing 40 mg dihydroartemisinin and 320 mg piperaquine | | |
| | Day 1 | Day 2 | Day 3 |
| 3 to 24 months | 0·5 | 0·5 | 0·5 |
| 2 to 7 years | 1 | 1 | 1 |
| 8 to 10 years | 1·5 | 1·5 | 1·5 |
| 11 to 14 years | 2 | 2 | 2 |
| 15+ years | 3 | 3 | 3 |
| **Primaquine (given on day 1)** | | | |
| Age | Number of tablets | Dosage (mg) | Volume (ml) of water used to dilute tablets |
| 2 to 4 years | 0·5 | 3·75 | 3 |
| 5 to 7 years | 0·75 | 5·625 | 5 |
| 8 to 10 years | 1 | 7·5 | 10 |
| 11 to 13 years | 1·5 | 11·25 | -- |
| 14+ years | 2 | 15 | -- |

**Seasonal malaria chemoprevention (control arm):**

| **Sulfadoxine-pyrimethamine + amodiaquine (SP-AQ)** | | | |
|---|---|---|---|
| Age | Number of tablets containing 500 mg sulfadoxine and 25 mg pyrimethamine, and 150 mg amodiaquine | | |
| | Day 1 | Day 2 | Day 3 |
| 3 to 11 months | 0·5 tablet SP + 0·5 tablet AQ | 0·5 tablet AQ | 0·5 tablet AQ |
| 12 to 59 months | 1 tablet SP + 1 tablet AQ | 1 tablet AQ | 1 tablet AQ |
| 60 to 120 months | 1·5 tablets SP + 1·5 tablets AQ | 1·5 tablets AQ | 1·5 tablets AQ |

**Appendix 3.** Boxplot of village-level distribution (A) and crude (B) coverage of mass drug administration (intervention) and seasonal malaria chemoprevention (control). Coverage is separately reported for overall population and for those under 10 years of age for comparability of MDA and SMC coverage.

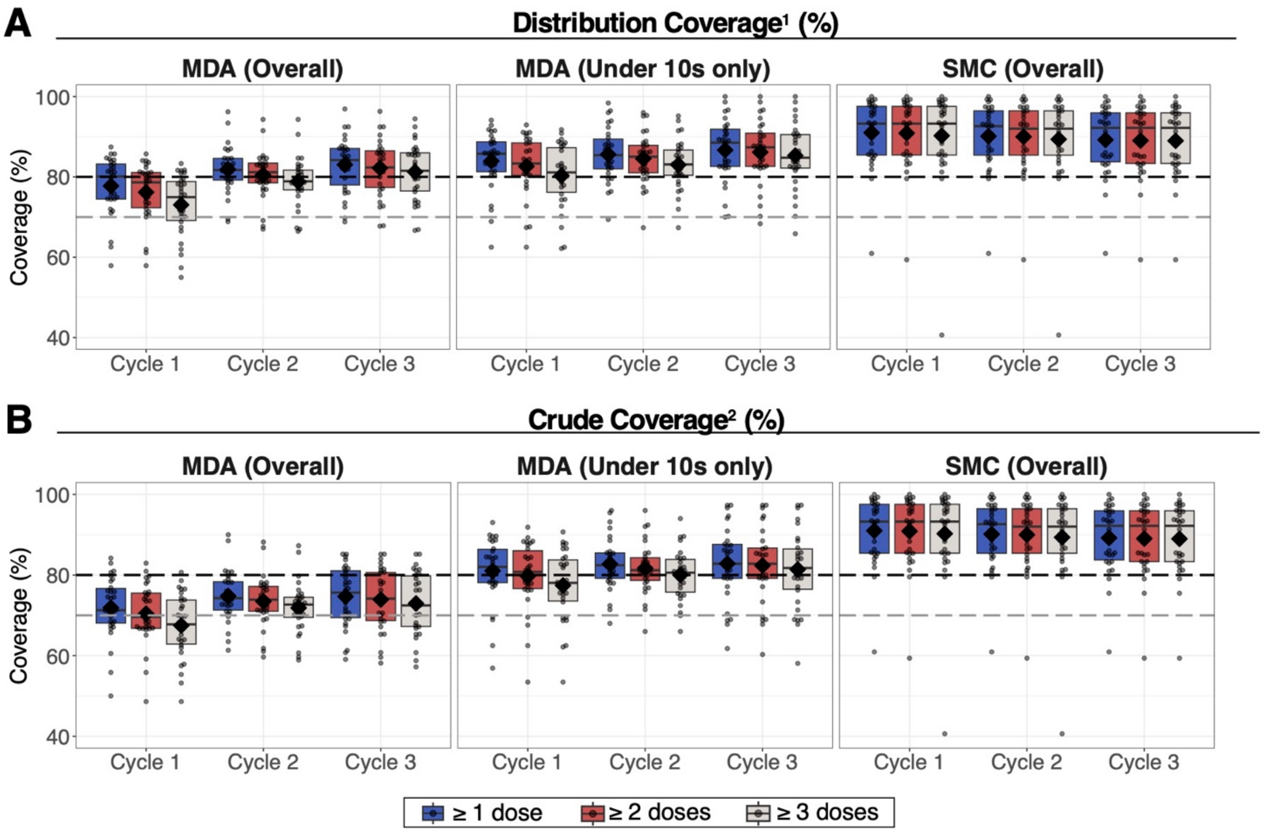


Abbreviations: MDA = mass drug administration; SMC = seasonal malaria chemoprevention

Boxes indicate the interquartile range of village-level coverage; horizontal line in each box indicates median; diamond point represents mean across villages; circle points indicate coverage for each village.

[1] Distributional coverage = # of residents who received study drugs / # of eligible residents targeted for campaign. Denominator includes absences and refusers of the study drugs, but excludes pregnant women (for MDA), persons with a self-reported illness, and persons who emigrated out of study area.

[2] Crude coverage = # of residents who received study drugs / # of persons targeted for the campaign. Denominator includes those who refused the study drugs, absences, and those who were ineligible to receive MDA due to pregnancy, self-reported illness, but excludes those who emigrated out of the study area.

**Appendix 4.** Reasons for non-coverage of MDA and SMC.

| **Intervention (MDA)** | **Cycle 1** | | | **Cycle 2** | | | **Cycle 3** | | |
|---|---|---|---|---|---|---|---|---|---|
| | **Day 1** | **Day 2** | **Day 3** | **Day 1** | **Day 2** | **Day 3** | **Day 1** | **Day 2** | **Day 3** |
| Targeted individuals[1] | 8931 | 8931 | 8931 | 9571 | 9571 | 9571 | 9703 | 9703 | 9703 |
| Received DP | 6449 (72·2%) | 6310 (70·7%) | 6163 (69·0%) | 7089 (74·1%) | 6986 (73·0%) | 6884 (71·9%) | 7237 (74·6%) | 7162 (73·8%) | 7085 (73·0%) |
| Received SLD-PQ[2] | 6286/8670 (72·5%) | -- | -- | 6949/9331 (74·5%) | -- | -- | 7199/9521 (75·6%) | -- | -- |
| Absent | 1649 (18·5%) | 1758 (19·7%) | 1888 (21·1%) | 1419 (14·8%) | 1482 (15·5%) | 1568 (16·4%) | 1311 (13·5%) | 1361 (14·0%) | 1423 (14·7%) |
| Refused | 121 (1·4%) | 125 (1·4%) | 129 (1·4%) | 146 (1·5%) | 161 (1·7%) | 169 (1·8%) | 131 (1·4%) | 135 (1·4%) | 140 (1·4%) |
| Self-reported illness | 458 (5·1%) | 484 (5·4%) | 497 (5·6%) | 613 (6·4%) | 638 (6·7%) | 646 (6·7%) | 701 (7·2%) | 722 (7·4%) | 732 (7·5%) |
| Pregnant/Breastfeeding | 254 (2·8%) | 254 (2·8%) | 254 (2·8%) | 304 (3·2%) | 304 (3·2%) | 304 (3·2%) | 323 (3·3%) | 323 (3·3%) | 323 (3·3%) |
| **Control (SMC)** | **Cycle 1** | | | **Cycle 2** | | | **Cycle 3** | | |
| | **Day 1** | **Day 2** | **Day 3** | **Day 1** | **Day 2** | **Day 3** | **Day 1** | **Day 2** | **Day 3** |
| Targeted individuals | 3492 | 3492 | 3492 | 3489 | 3489 | 3489 | 3487 | 3487 | 3487 |
| Received SP-AQ | 3202 (91·7%) | 3187 (91·3%) | 3199 (91·6%) | 3174 (91·0%) | 3159 (90·5%) | 3170 (90·9%) | 3146 (90·2%) | 3141 (90·1%) | 3139 (90·0%) |
| Absent | 234 (6·7%) | 248 (7·1%) | 236 (6·8%) | 247 (7·1%) | 261 (7·5%) | 249 (7·1%) | 261 (7·5%) | 261 (7·5%) | 262 (7·5%) |
| Refused | 21 (0·6%) | 22 (0·6%) | 22 (0·6%) | 21 (0·6%) | 22 (0·6%) | 22 (0·6%) | 27 (0·8%) | 28 (0·8%) | 28 (0·8%) |
| Self-reported illness | 35 (1·0%) | 35 (1·0%) | 35 (1·0%) | 47 (1·3%) | 47 (1·3%) | 48 (1·4%) | 53 (1·5%) | 57 (1·6%) | 58 (1·7%) |

Abbreviations: DP = dihydroartemisinin-piperaquine; MDA = mass drug administration; SLD-PQ = single low-dose primaquine; SMC = seasonal malaria chemoprevention

[1] Targeted individuals includes new residents that entered the study area, but excludes deaths and those who emigrated out of the study area.

[2] % calculated based on PQ eligibility (≥2 years of age).

**Appendix 5.** Safety outcomes reported through passive surveillance.

| Passive surveillance | Intervention (MDA) | Control (SMC) | p-value |
|---|---|---|---|
| Participants with any AE, n/N (%)[1] | | | |
| All cycles | 67/20 887 (0·003%) | 2/9524 (0·0002%) | < 0·0001 |
| First cycle | 46/6506 (0·7%) | 0/3202 (0%) | < 0·0001 |
| Second cycle | 13/7125 (0·2%) | 2/3174 (0·1%) | 0·14 |
| Third cycle | 8/7256 (0·1%) | 0/3146 (0%) | 0·062 |
| Frequency of adverse events, n (IR)[2] | | | |
| All cycles | 129 (0·0062) | 4 (0·00042) | < 0·0001 |
| First cycle | 97 (0·015) | 0 (0) | < 0·0001 |
| Second cycle | 20 (0·0028) | 4 (0·0013) | 0·13 |
| Third cycle | 12 (0·0017) | 0 (0) | 0·013 |
| Most common adverse events, n/N (%) | | | |
| Gastrointestinal issues | 59/129 (45·7%) | 3/4 (75%) | 0·25 |
| Headache | 33/129 (25·6%) | 0/4 (0%) | 0·24 |
| Fever | 22/129 (17·1%) | 1/4 (25%) | 0·37 |
| Other | 15/129 (11·6%) | 0/4 (0%) | 0·47 |
| Severity of AEs, n/N (%) | | | |
| Mild | 129/129 (100%) | 4/4 (100%) | -- |
| Moderate | 0/129 (0%) | 0/4 (0%) | -- |
| Severe | 0/129 (0%) | 0/4 (0%) | -- |
| Any serious AEs, n/N (%) | 0/129 (0%) | 0/4 (0%) | -- |

Abbreviations: AE = adverse event; IR = incidence rate

[1] Denominators represent those who received at least one dose of study drug.

[2] Incidence rate represents events per person-month at-risk. Denominators for incidence rate calculated based on number of persons who received at least one dose of study drug for the time of observation each person was monitored for adverse events.

**Appendix 6.** Prevalence of drug resistance markers.

| Mutations | Intervention | | Control | | p-value[1] |
|---|---|---|---|---|---|
| | **Baseline (n=128)** | **Endline (n=58)** | **Baseline (n=141)** | **Endline (n=106)** | |
| **PfK13** | | | | | |
| C580Y | 0% (0/123) | 0% (0/58) | 0% (0/135) | 0% (0/104) | -- |
| **PfDHPS** | | | | | |
| K540E | 0% (0/128) | 0% (0/58) | 0% (0/141) | 0% (0/106) | -- |
| A437G | 82% (96/117) | 87% (48/55) | 81% (103/127) | 87% (86/99) | 0·94 |
| A581G | 0% (0/128) | 0% (0/58) | 0% (0/141) | 0% (0/106) | -- |
| A613S/T | 14% (18/126) | 5% (3/57) | 16% (23/140) | 12% (12/101) | 0·33 |
| **PfDHFR** | | | | | |
| N51I | 100% (126/126) | 100% (56/56) | 100% (139/139) | 100% (105/105) | -- |
| C59R | 100% (126/126) | 100% (56/56) | 100% (139/139) | 100% (105/105) | -- |
| S108N | 75% (88/117) | 58% (32/55) | 77% (105/137) | 60% (61/102) | 0·96 |
| I164L | 12% (15/128) | 2% (1/54) | 12% (17/141) | 7% (1/101) | 0·24 |
| **PfCRT** | | | | | |
| K76T | 38% (48/127) | 35% (20/57) | 46% (63/138) | 44% (46/104) | 0·87 |
| **PfMDR1** | | | | | |
| N86Y | 2% (3/128) | 0% (0/58) | 5% (7/141) | 0% (0/106) | 0·28 |
| Y184F | 67% (80/120) | 64% (37/58) | 81% (107/132) | 64% (68/106) | 0·19 |

Abbreviations: Pf = *Plasmodium falciparum*; K13 = Kelch 13; DHPS = dihydropteroate synthase; DHFR = dihydrofolate reductase; CRT = chloroquine resistance transporter; MDR1 = multidrug resistance 1

[1] P-value computed using Poisson regression with robust standard errors, which compares relative change in prevalence before-and-after intervention between groups.

**Appendix 7.** Stratified analyses of intervention effectiveness on RDT-confirmed malaria incidence during peak transmission season (July-December).

| **Study outcomes** | **Average incidence per 1,000 (events / population)** | | | | **Intervention effect, 1-ratio of IRRs [95% CI][1]** | |
|---|---|---|---|---|---|---|
| | **Intervention (MDA)** | | **Control** | | | |
| | **2020** | **2021** | **2020** | **2021** | **Unadjusted** | **Adjusted[2]** |
| **DSDOM at baseline[3]** | | | | | | |
| Yes | 258 | 87 | 275 | 188 | 63% [41%, 77%] | 60% [34%, 76%] |
| No | 28 | 105 | 61 | 145 | -170% [-27%, -474%] | -15% [-289%, 66%] |

Abbreviations: IRR = incidence rate ratio; MDA = mass drug administration; SMC = seasonal malaria chemoprevention

[1] Intervention effect estimated using mixed effects Poisson regression with village-level random intercepts.

[2] Adjusted for DSDOM at baseline, village population size of children <10 years of age, distance from health post, baseline parasite prevalence by microscopy, and village population size in 2019.

[3] 20 out of 30 clusters in the intervention arm and 20 out of 30 clusters in the control arm contributed to the "Yes" category of the DSDOM at baseline analyses. Nineteen out of 20 (95%) clusters without DSDOM at baseline were defined as "Low" transmission intensity at baseline.
